# A Novel Scoring System for Early Assessment of the Risk of the COVID-19-associated Mortality in Hospitalized Patients: COVID-19 BURDEN

**DOI:** 10.1101/2022.01.09.22268975

**Authors:** Fatemeh Amirzadehfard, Mohammad Hossein Imanieh, Sina Zoghi, Faezeh Sehatpour, Peyman Jafari, Hamidreza Hassanipour, Maryam Feili, Maryam Mollaie, Pardis Bostanian, Samrad Mehrabi, Reyhaneh Dashtianeh, Afrooz Feili

## Abstract

**Background:** Corona Virus Disease 2019 (COVID-19) presentation resembles common flu or can be more severe; it can result in hospitalization with significant morbidity and/or mortality. We made an attempt to develop a predictive model and a scoring system to improve the diagnostic efficiency for COVID-19 mortality via analysis of clinical features and laboratory data on admission.

**Methods:** We retrospectively enrolled 480 consecutive adult patients, aged 21-95, who were admitted to Faghihi Teaching Hospital. Clinical and laboratory features were extracted from the medical records and analyzed using multiple logistic regression analysis.

**Results:** A novel mortality risk score (COVID-19 BURDEN) was calculated, incorporating risk factors from this cohort. CRP (> 73.1 mg/L), O2 saturation variation (greater than 90%, 84-90%, and less than 84%), increased PT (>16.2s), diastolic blood pressure (≤75 mmHg), BUN (>23 mg/dL), and raised LDH (>731 U/L) are the features comprising the scoring system. The patients are triaged to the groups of low- (score <4) and high-risk (score ≥ 4) groups. The area under the curve, sensitivity, and specificity for predicting non-response to medical therapy with scores of ≥ 4 were 0.831, 78.12%, and 70.95%, respectively.

**Conclusion:** Using this scoring system in COVID-19 patients, the severity of the disease will be determined in the early stages of the disease, which will help to reduce hospital care costs and improve its quality and outcome.

## Introduction

Corona Virus Disease 2019 (COVID-19) caused by the Severe Acute Respiratory Syndrome Coronavirus-2 (SARS-CoV-2) is highly contagious. Symptoms of COVID-19 can be very similar to the common flu that include fever, cough, and congestion of the nasal cavity (1). As the pandemic spread, other symptoms such as loss of taste and smell (anosmia) have also emerged (2, 3). Patients with the severe form of the disease can experience a large range of symptoms arising from the host immune response including serious respiratory disease and pneumonia, vascular and hemodynamic disorders, and metabolic dysfunction (4-7). Patients can also present normal or abnormal leukocyte counts, lymphopenia, or thrombocytopenia, with extended activated thromboplastin time and increased C-reactive protein (CRP) level (8-10). Those most at risk are the elderly and people with medical comorbidities, such as cardiovascular disorders and diabetes mellitus (11-13). The exact mechanisms behind the disease and why some remain asymptomatic carriers while other patients develop severe disease with the fatal outcome are still poorly understood (14).

Here, we present details of the patients with laboratory-confirmed COVID-19 pneumonia to shed light on the characteristics of patients who experienced in-hospital mortality and explore the risk factors that might facilitate early screening. We develop a predictive model and scoring system to improve the diagnostic efficiency for COVID-19 mortality via analysis of clinical features and laboratory data on admission, allowing timely intervention.

## Materials and Methods

We retrospectively investigated 480 consecutive adult patients, aged 21–95, who were admitted to Faghihi Teaching Hospital from September 23, 2020, to November 21, 2020, the main referral center for COVID-19 in Shiraz, Fars province, Iran. The patients who had a positive RT-PCR COVID-19 test and were admitted during the study were included in this study. The patients who did not complete their course of hospitalization and were released with their consent were excluded. The study protocol was approved by the local ethics committee of Shiraz University of Medical Sciences (IR.SUMS.MED.REC.1400.382). Written informed consents were obtained from all participants on admission.

Corresponding medical records were thoroughly examined, retrospectively. Variables investigated in this study for correlation with the outcome of hospitalization (discharge or death) included demographic variables (gender and age), previous medical conditions (smoking/opium abuse, hypertension, HTN, and diabetes mellitus, DM, ischemic heart disease, IHD, and hyperlipidemia, and HLP), previous drug history (angiotensin-converting-enzyme inhibitors/angiotensin II receptor blockers; ACEIs/ARBs, calcium channel blocker; CCB, beta blockers; BB, acetylsalicylic acid; ASA, and Statins) the patients’ condition on admission (systolic blood pressure; SBP, diastolic blood pressure; DBP, pulse rate; PR, respiratory rate; RR, temperature; T, O2 saturation; O2 Sat, presence of dyspnea, cough, chest pain, fever, malaise, anorexia, nausea and/or vomiting; N/V, other gastrointestinal symptoms, and the interval between disease onset and hospital admission), the laboratory findings of the sample obtained at admission (white blood cell count; WBC, absolute neutrophil count; ANC, absolute lymphocyte count; ALC, hemoglobin; Hb, prothrombin time; PT, partial thromboplastin time; PTT, blood urea nitrogen; BUN, creatinine; Cr, sodium; Na, potassium; K, aspartate transaminase; AST, alanine transaminase; ALT, alkaline phosphatase; ALP, albumin; Alb, total bilirubin; TB, direct bilirubin; DB, creatinine phosphokinase; CPK, lactate dehydrogenase; LDH, erythrocyte sedimentation rate; ESR, C-reactive protein; CRP), and total length of stay in the hospital.

### Statistical Analysis

Quantitative variables are presented as mean ± SD and qualitative variables as frequency (percentage) for all independent variables. Univariate analysis was applied to identify the potential risk factors of mortality through independent t-test and chi-square test as appropriate. Statistically significant variables (significance level set at 0.05) that were of concern from a clinical perspective were subsequently employed in multiple logistic regression analysis with backward elimination method to identify the predictive factors. To obtain a clinical predictive risk score, we categorized the continuous prognostic variables using receiver operating characteristic (ROC) curve. The resulting variables were re-entered into a logistic regression model. According to relative contribution of each variable in the logistic regression model, which was determined by the regression coefficient, an integer score was assigned to each categorical variable (15). Lastly, the scores were arranged to obtain a practical triage of the patients into low- and high-risk cases. SPSS version 23 (SPSS Inc, IBM, New York, NY) and MedCalc statistical program, version 19.5 (MedCalc Software, Mariakerke, Belgium) were used to analyze the data.

## Results

### Patient Details and Hospitalization

A total of 1511 patients were visited at the triage of Faghihi hospital with the impressions related to COVID-19, of whom 480 patients (212 females and 268 males) fulfilled the criteria set by this study. The median age of the patients included was 61 years old (IQR: 49-72). The median days from the onset of symptoms to admission was 8±6 days, and total length of stay in hospital was 5 days (IQR: 3-9). Intensive care unit (ICU) admission of COVID-19 patients is significantly associated with the worse outcome of hospitalization (P-value <0.0001).

### Comparison of the Associated Factors

The patients were assigned into two groups (discharged = 312 and expired = 168). In total, 48 demographic, clinical, and laboratory variables were compared in both groups according to the outcome of hospitalization to determine meaningful ones (Table S1). Multiple logistic regression analysis was carried out separately to assess the effect of 19 variables identified as significant independent predictors of outcome in our cohort (Table 1).

**Table 1.** Demographic, clinical, and laboratory characteristics of COVID-19 patients significantly associated with mortality, stratified by the outcome of hospitalization. These variables entered multivariate logistic regression analysis. HTN, hypertension; IHD, ischemic heart disease; N/V, nausea and/or vomiting; BP, blood pressure; RR, respiratory rate; O2 Sat, O2 saturation; WBC, white blood cell count; ANC, absolute neutrophil count; PT, prothrombin time; BUN, blood urea nitrogen; Cr, creatinine; K, potassium; AST, aspartate transaminase; Alb, albumin; CPK, creatinine phosphokinase; LDH, lactate dehydrogenase; CRP, C-reactive protein.

| Variable | Discharged (n =312) | Expired (n =168) | *P value* | Cut-off set for Ratio Variables |
| --- | --- | --- | --- | --- |
| Age<br>Mean (SD) | 57.03 (15.759) | 66.80 (13.930) | *<0.0001* | <60<br>≥60 |
| HTN<br>(Present:<br>Absent) | 115: 197 | 80:88 | *0.025* | - |
| IHD<br>(Present:<br>Absent) | 43: 269 | 40: 128 | *0.008* | - |
| N/V<br>(Present:<br>Absent) | 75: 236 | 27: 140 | *0.047* | - |
| Systolic BP | 127.97 (19.083) | 123.05 (24.884) | *0.026* | >114<br>≤114 |
| Diastolic BP | 79.44<br>(11.350) | 75.06 (13.180) | *0.001* | >75<br>≤75 |
| RR | 20.50 (4.715) | 23.86 (7.436) | *<0.0001* | ≤20<br>>20 |
| O<sub>2</sub> Sat | 84.63 (10.539) | 70.42 (16.926) | *<0.0001* | >90<br>83-90<br>≤83 |
| WBC | 7.7684 (4.17933) | 10.1776 (5.87533) | *<0.0001* | ≤7.52<br>>7.52 |
| ANC | 6045.06<br>(3850.1621) | 8573.50 (5173.281) | *<0.0001* | >5628<br>≤5627 |
| PT | 32.92 (23.700) | 42.31 (26.991) | *<0.0001* | ≤16.2<br>>16.2 |
| BUN | 21.16 (26.450) | 34.76 (24.786) | *<0.0001* | <=23<br>>23 |
| Cr | 1.4259 (1.46007) | 1.9012 (1.81179) | *0.002* | <=1.3<br>>1.3 |
| K | 4.452 (0.5787) | 4.811 (0.8522) | *<0.0001* | <=4.9<br>>4.9 |
| AST | 67.74 (99.519) | 116.94 (305.125) | *0.015* | <=67<br>>67 |
| Alb | 3.748 (0.3469) | 3.446 (0.4644) | *<0.0001* | >3.7<br><=3.7 |
| CPK | 218.35 (312.488) | 634.76 (1432.796) | *<0.0001* | <=78<br>>78 |
| LDH | 742.83 (561.077) | 1055.28 (637.258) | *<0.0001* | <=731<br>>731 |
| CRP | 64.39 (50.420) | 72.99 (21.225) | *0.049* | <=73.1<br>>73.1 |

To generate a scoring system, O2 Sat, DBP, PT, BUN, LDH, and CRP values, among other continuous variables, were categorized using integer cut-points guided by the receiver– operator characteristic (ROC) curve and observed relationship with outcomes.

### Final Model and COVID-19 BURDEN Risk Score

To formulate a numerical scoring, we used the coefficients generated by the logistic regression equation, to create an integer number, approximating the values of the coefficients for each of the categories in Table 2.

**Table 2.** The variables, which remained in the final multiple logistic regression model to predict the risk of mortality during hospitalization. O2 Sat >90% was used as reference index in O2 saturation category. Formulation of integer risk score for each category was based on the strength of contribution to logistic equation based on the coefficient (for example, the coefficient of O2 Sat <84 % is 2.218; therefore, an integer score of 2 was given. Coefficient of BUN is 1.254; therefore, an integer score of 1 is given). DBP, diastolic blood pressure; PT, prothrombin time; BUN, blood urea nitrogen; LDH, lactate dehydrogenase; CRP, C-reactive protein; O2 Sat, O2 saturation.

| Variables | Coefficient | Coefficient (SE) | *P value* | Odds ratio (95% CI) | Lower – Upper 95% CI |
| --- | --- | --- | --- | --- | --- |
| DBP | 0.794 | 0.360 | *0.027* | 2.212 | 1.093-4.479 |
| PT | 0.761 | 0.349 | *0.029* | 2.140 | 1.079-4.242 |
| BUN | 1.254 | 0.356 | *<0.0001* | 3.503 | 1.744-7.039 |
| LDH | 1.066 | 0.369 | *0.004* | 2.903 | 1.408-5.987 |
| CRP | 1.017 | 0.418 | *0.015* | 2.766 | 1.220-6.271 |
| 84% ≤ O2 Sat ≤90% | 0.799 | 0.652 | *<0.0001* | 2.223 | 0.620-7.975 |
| O2 Sat <84 % | 2.218 | 0.604 | *<0.0001* | 9.189 | 2.811-30.040 |

Using the coefficients of the regression analyses, the O2 Sat greater than 90%, 84-90%, and less than 84% were given scores of 0, 1, and 2, respectively. Scores of either 0 or 1 were given to other variables including CRP, PT, DBP, BUN, and LDH levels (Table 3).

**Table 3.** Integer risk score attributable to each category derived from the coefficients of the logistic regression equation. The overall COVID-19 BURDEN risk score is between 0 and 7; the higher the score, the higher the risk of mortality during the course of hospitalization. The cut-off of the risk factor is set at 4. The patients with a score <4 are categorized as low-risk, having a more favorable outcome, while those with a score ≥4 were more likely to have an undesirable outcome.

| Variables | Points Awarded |
| --- | --- |
| **C-reactive protein (CRP)** |  |
| ≤ 73.1 mg/L | 0 |
| > 73.1 mg/L | 1 |
| **O<sub>2</sub> saturation Variation (O<sub>2</sub> sat Variation)** |  |
| Greater than 90% | 0 |
| 84-90% | 1 |
| Less than 84% | 2 |
| **Increased prothrombin time (Increased PT)** |  |
| ≤ 16.2 s | 0 |
| > 16.2 s | 1 |
| **Diastolic blood pressure (DBP)** |  |
| >75 mmHg | 0 |
| ≤ 75 mmHg | 1 |
| **Blood Urea Nitrogen (BUN)** |  |
| ≤ 23 mg/dL | 0 |
| > 23 mg/dL | 1 |
| **Raised Lactate Dehydrogenase (Raised LDH)** |  |
| ≤ 731 Units/Lit (U/L) | 0 |
| > 731 Units/Lit (U/L) | 1 |
| **COVID-19 BURDEN risk score = (CRP score) + (O<sub>2</sub> sat Variation score) + (Increased PT score) + (DBP score) + (BUN score) + (Raised LDH score)** |  |

A novel mortality risk score (termed COVID-19 BURDEN) was calculated, incorporating risk factors from this cohort. COVID-19 BURDEN is an acronym for **C**RP (> 73.1 mg/L), **O**2 saturation Variation (greater than 90%, 84-90%, and less than 84%), **I**ncreased PT (>16.2s), **D**iastolic blood pressure (≤75 mmHg), **BU**N (>23 mg/dL), and **R**aised LDH (>731 U/L), as shown in Table 3.

Using the new scoring system, we scored each patient and compared him/her with his/her eventual outcomes. For an individual patient, the total score was derived from the sum of the score attributed to the variables mentioned above. The minimum score possible was 0 while the maximum was 7.

It was possible to triage the patients to the groups of low- (score <4) and high-risk (score ≥ 4) groups. ROC analysis for this risk score yielded an area under the curve (AUC) of 0.831 (AUC of 1.0 indicating a perfect test) (Figure 1). The sensitivity and specificity for predicting non-response to medical therapy with scores of ≥ 4 were 78.12% and 70.95%, respectively.

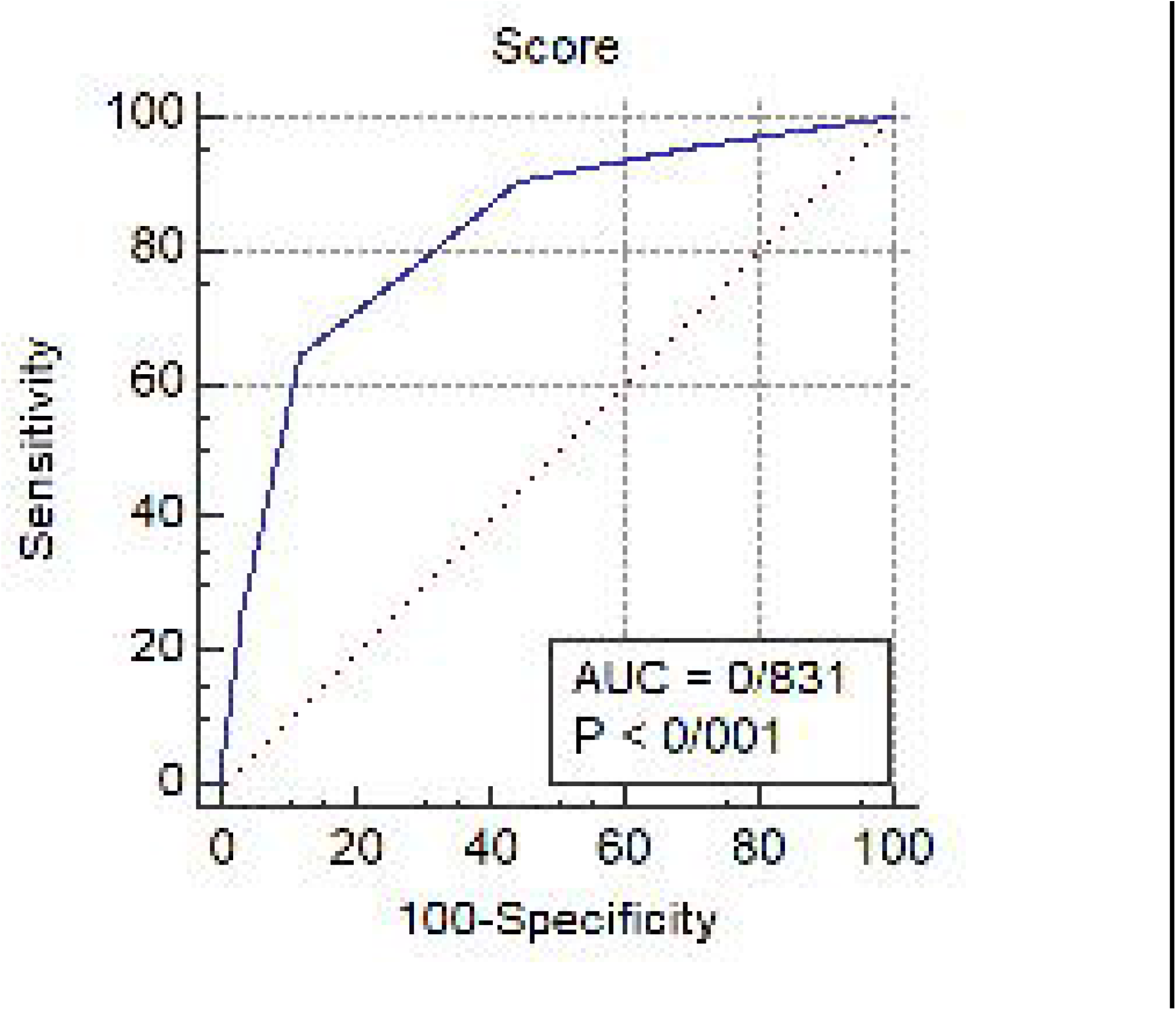

## Discussion

COVID-19 pandemic is a public health concern with dire health, environmental, and economic consequences (16). Currently, the facilities required for the hospitalization of patients with Covid-19 are limited. This limitation is especially pronounced in ICUs for patients requiring mechanical ventilation (17). It is necessary to develop a scoring system that can be used by physicians, to estimate the severity of the disease and prognosis of each patient at an early stage of the disease, which will grant a wider timespan for interventions. This tool also helps to reduce hospital care costs and improves its quality in the health care units (18-20).

In this survey, we designed a simplly calculated clinical risk score, by using the patient’s clinical characteristics and laboratory data, which can be used as a predictor to estimate the severity of the disease, the need for hospitalization, and the possibility of ICU admission requirement during hospitalization.

Low DBP (≤ 75 mmHg), prolonged PT (> 16.2s), increase in the level of LDH (> 731 U/L), BUN >23mg/dl, elevated CRP (> 73.1 mg/L), and decrease in oxygen saturation (<84%) were detected as risk factors for disease severity among 480 adult patients. Using this scoring system is a useful strategy for screening high-risk patients in crowded care centers during the COVID-19 outbreak.

Severe hypoxemia (O2 sat<84%) had the highest odds ratio (OR= 9.19) among the 6 risk factors determining the severity of the disease in our survey. This finding was consistent with a cohort study by Bahl et al. among 1461 patients in which O2 sat of ≤188% was associated with a higher mortality rate (21). In the review conducted by Petrilli et al. among 4103 patients, oxygen saturation of < 88% (OR= 6.99) was also introduced as the most critical predicting factor on admission (22).

Recently, several studies have been conducted in different parts of the world to develop a simple scoring system to predict the prognosis and outcome of the COVID-19 (20, 23-26). The first scoring system to predict the severity of COVID-19, incorporating age, glomerular filtration rate (GFR), WBC, neutrophil count, and myoglobin, was developed by Zhang et al. in 2020 among 80 patients (23). A low sample size was one of the limitations of their study. Moreover, the afore-mentioned scoring system did not include any of the vital signs, which are quite important in determining the clinical course of COVID-19 patients. The findings of the study conducted by Altschul et al. are also in agreement with the present study. Old age (especially ≥ 80 years), mean arterial blood pressure≤ 60 mmHg, O2 sat <94%, BUN>30mg/dl, CRP>10 mg/dl, and INR >1.2 were identified as six risk factors that affect the mortality rate of COVID-19 patients. Their study was conducted in three major referral hospitals of New York City. However, the study data is limited to their urban population and may not be fully generalizable to all countries and races (25). In another study which was published in July 2020, old age, presence of coronary heart disease, high procalcitonin, lymphopenia, and high d-Dimer level were associated with a high mortality rate in the affected population (26).

Having a precise, inexpensive, accessible, and straightforward scoring system for all health care providers will improve the patients’ triage, and help to identify high-risk individuals, initiate timely interventions, and allocate resources efficiently (23, 27). One of the advantages of our scoring system is that it doesn’t include lung high resolution computed tomography (HRCT) scans, which contributes greatly to the health financial system in this epidemic. Rather, we used more accessible clinical and laboratory metrics to predict disease severity, which will reduce the costs, improve the timing of the decision making in the initial screening, and lift the burden from the already exhausted imaging facilities.

CRP, Oxygen saturation variation, increased PT, DBP and BUN, and raised LDH (COVID-19 BURDEN) were detected as six factors for COVID-19 infection severity. Using this scoring system in COVID-19 patients, the severity of the disease in the early stages of the disease can be estimated and helps to reduce health costs and improve the quality of patient care in the health care units.

One of the limitations of this study was that this study was performed only in one center of COVID-19 patients although this center is one of the tertiary referral centers in the south of the country. However, further studies with larger sample size are needed in different areas and among different ethnicities and races. For a more accurate and comprehensive summary, more investigation with a larger database in different regions, especially in developing countries, is needed.

## Data Availability

All data produced in the present work are contained in the manuscript.

## Declarations

### Ethical Approval

The study protocol was approved by the local ethics committee of Shiraz University of Medical Sciences (IR.SUMS.MED.REC.1400.382). Written informed consents were obtained from all participants on admission.

## Acknowledgements

The authors would like to thank Shiraz University of Medical Sciences, Shiraz, Iran and also Center for Development of Clinical Research of Nemazee Hospital and Dr. Nasrin Shokrpour for editorial assistance.

## Funding

The authors received no financial support for the research, authorship, or publication of this article.

## Availability of data and material

All data generated or analyzed during this study are included in the final published article.

## Notes

### Competing Interest Statement

The authors have declared no competing interest.

### Funding Statement

This study did not receive any funding.

### Author Declarations

Ethics committee/ of Shiraz University of Medical Sciences gave ethical approval for this work.

