## Supplemental Table 1 for "A Novel Scoring System for Early Assessment of the Risk of the COVID-19-associated Mortality in Hospitalized Patients: COVID-19 BURDEN"

| Variable | Discharged (n =312) | Expired (n =168) | *P value* |
| --- | --- | --- | --- |
| Age  Mean (SD) | 57.3 (15.759) | 66.80 (13.930) | ***<0.0001*** |
| Gender (Male: Female) | 170:142 | 98:70 | *0.442* |
| Smoking/ Opium (Smoker: Non-smoker) | 16:296 | 13:155 | *0.315* |
| HTN (Present: Absent) | 115: 197 | 80:88 | ***0.025*** |
| DM (Present: Absent) | 87: 225 | 52: 116 | *0.527* |
| IHD (Present: Absent) | 43: 269 | 40: 128 | ***0.008*** |
| HLP (Present: Absent) | 40: 272 | 24: 144 | *0.674* |
| Dyspnea (Present: Absent) | 216: 96 | 125: 43 | *0.243* |
| Cough (Present: Absent) | 188: 124 | 89: 79 | *0.145* |
| Chest pain (Present: Absent) | 28: 284 | 19: 149 | *0.423* |
| Fever (Present: Absent) | 174: 138 | 90: 78 | *0.700* |
| Malaise (Present: Absent) | 163: 149 | 80: 88 | *0.339* |
| Anorexia (Present: Absent) | 64: 248 | 36: 132 | *0.815* |
| N/V (Present: Absent) | 75: 237 | 27: 141 | ***0.047*** |
| H/A (Present: Absent) | 29: 283 | 10:158 | *0.225* |
| GI symptoms (Present: Absent) | 52: 260 | 33: 135 | *0.451* |
| ACEI/ARB (Received: Not received) | 51: 261 | 31: 137 | *0.611* |
| CCB (Received: Not received) | 26: 282 | 19: 149 | *0.327* |
| BB (Received: Not received) | 18: 292 | 17: 151 | *0.098* |
| ASA (Received: Not received) | 48: 262 | 27: 139 | *0.895* |
| Statin (Received: Not received) | 39: 270 | 29: 138 | *0.171* |
| Systolic BP | 127.97 (19.083) | 123.05 (24.884) | ***<0.0001*** |
| Diastolic BP | 79.44 (11.350) | 75.06 (13.180) | ***0.026*** |
| PR | 92.77 (13.567) | 92.52 (17.54) | *0.874* |
| RR | 20.50 (4.715) | 23.86 (7.436) | ***<0.0001*** |
| T | 36.879 (0.7688) | 36.936 (0.7594) | *0.483* |
| O_2_ Sat | 84.63 (10.539) | 70.42 (16.926) | ***<0.0001*** |
| Onset To Admission | 8.17 (5.107) | 7.85 (7.433) | *0.617* |
| WBC | 7.7684 (4.17933) | 10.1776 (5.87533) | ***<0.0001*** |
| ANC | 6045.06 (3850.1621) | 8573.50 (5173.281) | ***<0.0001*** |
| ALC | 1195.71 (737.939) | 1089.36 (860.409) | *0.167* |
| Hb | 12.708 (1.9789) | 12.387 (2.3133) | *0.119* |
| PT | 32.92 (23.700) | 42.31 (26.991) | ***<0.0001*** |
| PTT | 34.43 (13.102) | 33.76 (9.930) | *0.577* |
| BUN | 21.16 (26.450) | 34.76 (24.786) | ***<0.0001*** |
| Cr | 1.4259 (1.46007) | 1.9012 (1.81179) | ***0.002*** |
| Na | 139.47 (3.947) | 138.77 (5.159) | *0.103* |
| K | 4.452 (0.5787) | 4.811 (0.8522) | ***<0.0001*** |
| AST | 67.74 (99.519) | 116.94 (305.125) | ***0.015*** |
| ALT | 58.31 (85.921) | 91.42 (357.332) | *0.144* |
| ALP | 240.60 (138.879) | 263.34 (198.614) | *0.171* |
| Alb | 3.748 (0.3469) | 3.446 (0.4644) | ***<0.0001*** |
| TB | 0.8767 (0.62512) | 1.0025 (0.79613) | *0.070* |
| DB | 0.3197 (0.40510) | 0.4082 (0.54844) | *0.057* |
| CPK | 218.35 (312.488) | 634.76 (1432.796) | ***<0.0001*** |
| LDH | 742.83 (561.077) | 1055.28 (637.258) | ***<0.0001*** |
| ESR | 57.39 (31.486) | 54.76 (31.019) | *0.474* |
| CRP | 64.39 (50.420) | 72.99 (21.225) | ***0.049*** |

Table S1. Demographic, clinical, and laboratory characteristics of COVID-19 patients,
stratified by the outcome of hospitalization
